# Antimicrobial resistance determinants are associated with *Staphylococcus aureus* bacteraemia and adaptation to the hospital environment: a bacterial genome-wide association study

**DOI:** 10.1101/2021.01.13.21249734

**Authors:** Bernadette C Young, Chieh-Hsi Wu, Jane Charlesworth, Sarah Earle, James R Price, N Claire Gordon, Kevin Cole, Laura Dunn, Elian Liu, Sarah Oakley, Heather Godwin, Rowena Fung, Ruth Miller, Kyle Knox, Antonia Votintseva, T Phuong Quan, Robert Tilley, Matthew Scarborough, Derrick W Crook, Timothy E Peto, A Sarah Walker, Martin J Llewelyn, Daniel J Wilson

## Abstract

**Background:** *Staphylococcus aureus* is a major bacterial pathogen in humans, and a dominant cause of severe bloodstream infections. Globally, antimicrobial resistance (AMR) in *S. aureus* remains challenging. While human risk factors for infection have been defined, contradictory evidence exists for the role of bacterial genomic variation in *S. aureus* disease.

**Methods:** To investigate the contribution of bacterial lineage and genomic variation to the development of bloodstream infection, we undertook a genome-wide association study comparing bacteria from 1017 individuals with bacteraemia to 984 adults with asymptomatic *S. aureus* nasal carriage. Within 984 carriage isolates, we also compared healthcare-associated (HA) carriage with community-associated (CA) carriage.

**Results:** All major global lineages were represented in both bacteraemia and carriage, with no evidence for different attack rates. However, kmers tagging trimethoprim resistance-conferring mutation F99Y in *dfrB* were significantly associated with bacteraemia-vs-carriage (*p=*10^−8.9^-10^−9.3^). Pooling variation within genes, bacteraemia-vs-carriage was associated with the presence of *mecA* (HMP=10^−5.3^) as well as the presence of SCCmec (HMP=10^−4.4^).

Among *S. aureus* carriers, no lineages were associated with HA-vs-CA carriage. However, we found a novel signal of HA-vs-CA carriage in the foldase protein *prsA*, where kmers representing conserved sequence allele were associated with CA carriage (*p=*10^−7.1^-10^−19.4^), while in *gyrA*, a ciprofloxacin resistance-conferring mutation, L84S, was associated with HA carriage (*p=*10^−7.2^).

**Conclusions:** In an extensive study of *S. aureus* bacteraemia and nasal carriage in the UK, we found strong evidence that all *S. aureus* lineages are equally capable of causing bloodstream infection, and of being carried in the healthcare environment.

Genomic variation in the foldase protein *prsA* is a novel genomic marker of healthcare origin in *S. aureus* but was not associated with bacteraemia. AMR determinants were associated with both bacteraemia and hospital-associated carriage, suggesting that AMR increases the propensity not only to survive in hospital environments, but also to cause invasive disease.

## Background

*Staphylococcus aureus* is a common coloniser of human mucosal surfaces and skin but also a major human pathogen. ^1,2^ It is a leading cause of hospital and community acquired infection and one of the leading causes of bloodstream infection worldwide.^1,2,3,4^ Over 12,000 cases occur each year in England, and the rate continues to increase, despite improving control of Methicillin-resistant *S. aureus* (MRSA) bacteraemia.^5^ Mortality following *S. aureus* bacteraemia (SAB) has not declined over recent years, and remains generally over 20% at 30 days, even with appropriate antimicrobial therapy.^6,7,8^

*S. aureus* possesses diverse and variable virulence mechanisms facilitating tissue invasion, inflammation and evasion of host immune factors. These include a thick peptidoglycan wall, polysaccharide capsule,^1^ toxins,^9,10^ complement control proteins,^11^ and bound adhesins.^12^ With the exception of specific toxinoses such as Toxic Shock Syndrome,^13,14,15^ and the role of Panton-Valentine Leucocidin (PVL) in skin or soft tissue infections^16^ and pyomyositis,^17^ evidence linking bacterial genetic variability to clinical disease phenotype is inconclusive.

The majority of evidence accrues from case-control studies using candidate gene approaches or microarrays to examine gene presence or absence. Such studies have implicated several specific genes encoding putative virulence factors in invasive *S. aureus* disease. Secreted enterotoxins,^18,19,20,21^ haemolysins^18^ and leucotoxins^22^; surface proteins which mediate tissue attachment, invasion and immune evasion^18,22^, the presence of the intercellular adhesin locus^18,23^ and variation in the Agr regulatory system^22^ have all been shown to co-occur with invasive *S. aureus*. However, the evidence for these associations is inconsistent, and for every study reporting an association, there is at least one large study shows no evidence of effect.^18,24^

The fact that a few *S. aureus* lineages account for the majority of *S. aureus* infections suggest important inter-lineage differences in virulence but again evidence is conflicting for a relationship between Clonal Complex (CC) and invasive disease.^22,24,25,26,27,28,29^ These discrepant results may reflect the variable sensitivity of probes employed, or the inconsistent methods used to control for effects of population structure. In particular, associations cannot be reliably inferred without considering linkage disequilibrium between candidate genes and potential virulence factors elsewhere in the genome.

Different genomic associations have been identified for community-associated (CA) or healthcare-associated (HA) SAB.^30^ Loss of function mutations in the Accessory Gene Regulator (Agr), a central controller of *S aureus* expression, have been frequently found in HA bloodstream infections,.^31^ Reduced cytotoxicity and low Agr expression were also independent predictors of mortality in a study of nosocomial MRSA pneumonia.^32^ There is a direct relationship between MecA expression and Agr dysfunction: the altered Penicillin Binding Protein (PBP) expressed by some MRSA can directly reduce Agr mediated toxin production.^33^ This dysfunction may be a fitness ‘cost’, overcome by the relative advantage of antimicrobial resistance in healthcare settings, and the relatively lower host defences found in hospitalised patients. However, evidence shows MRSA clones traditionally conceived of as HA lineages – such as clonal complex (CC) 22 - are equally capable of transmission in the community, including household settings,^34^ questioning the notion that MRSA requires the healthcare setting to gain a relative advantage.

There is growing evidence that the relationship between toxin production and virulence in *S. aureus* is not straightforward. While superantigen toxins and leukocidins have been linked to certain disease phenotypes, genomic changes associated with reduced bacterial toxicity may actually enhance bacterial survival in the bloodstream, evidenced by lower lymphocyte toxicity and greater fitness in human serum exhibited by bacteraemia isolates compared to those found in nasal carriage or soft tissue infection.^42^ Agr defective strains have been found in association with persistent bacteraemia,^36,37^ and associated with higher mortality.^38^Naturally occurring loss of function mutations in the regulatory protein Repressor of Surface Proteins (Rsp) have been documented arising within-host and in bloodstream infections.^39^ These mutations were associated with attenuated mortality in a mouse model of disease, but preserved the ability to disseminate and form abscesses,^40^ and have been shown to alter survival in blood and antibiotic tolerance.^41^ A comprehensive survey of within-host evolution of *S. aureus* infection demonstrated evidence of bacterial genomic adaption, with protein altering variation in regulatory genes, and the cell surface proteins under control of key regulators.^43^ Similar signals of adaptation were found in the genetic changes associated with prolonged bacteraemia.^44^

Thus, while conflicting evidence for the role of gene presence in carriage-vs-bacteraemia arises from case-control studies, there are observations supporting the hypothesis that subtle genetic variation – including that of a type traditionally thought to diminish virulence – could increase the likelihood of SAB. Recent developments in bacterial genome wide association studies (GWAS) demonstrate that these powerful tools can help delineate the genomic basis of bacterial infection. A study of bacteraemia caused by the ST239 lineage investigated associations between bacterial genetic variants, toxin production and severity of disease in a mouse model.^45^ Genomic variants were integrated with bacterial phenotyping and clinical data in 300 adults with bacteraemia involving the CC22 and CC30 lineages, finding that bacterial predictors of mortality varied by lineage.^46^ Investigating whether a genomic basis for invasive disease exists more generally at a population level requires careful control for population structure, an otherwise potent confounder. Bacterial GWAS incorporating such controls has recently identified PVL as the key determinant of *S. aureus* pyomyositis in a paediatric population.^17^

Here we present a bacterial GWAS of SAB across bacterial lineages, studying population-representative cases of bacteraemia and nasal carriage controls, integrating clinical data with 2001 bacterial sequences to investigate whether bacterial lineage or genomic variation is associated with carriage-vs-bacteraemia. Within *S. aureus* carriage, we further examine genomic features associated with CA-vs-HA carriage.

## Materials and methods

### Identification of cases and controls

Cases of *S. aureus* bacteraemia (SAB) were identified from 3 UK hospital trusts between 2008-2014: Oxford University Hospitals NHS trust (Oxford UK), Brighton and Sussex University Hospitals NHS trust (Brighton, UK) and University Hospitals Plymouth NHS trust (Plymouth, UK). These sites were part of the UK Clinical Infection Research Group (UKCIRG) which established prospective cohort study of SAB in 2008,^6^ and then the International *Staphylococcus Aureus* Collaboration (ISAC) which established a multinational prospective cohort study of SAB in 2006.^7^ Sequential individuals from these studies over 13 years of age with *S. aureus* on blood culture were included if there was an isolate available for sequencing, with associated clinical data, and the blood culture had not been deemed to be a contaminant on local clinical review. We identified 1203 cases in patients that were not contaminants, after excluding repeat episodes (775, 232 and 196 at each centre respectively). Bacterial isolates were found from 724, 207 and 163 episodes at each centre respectively, and a minimum clinical data set (see below) was available for 674, 187 and 160 cases. We successfully sequenced 1017 of these 1021 cases for inclusion.

These included 417 cases from a previously sequenced collection investigating identifying antimicrobial resistance.^47^

*S. aureus* isolates from nasal carriage in individuals without *S. aureus* infection were identified from two studies of *S. aureus* carriage in Oxfordshire, UK (Figure S2).

The first was a study of *S. aureus* nasal carriage in adults in the community between July 2009 and April 2013. ^48^ Of 1123 individuals enrolled, 360 individuals carrying *S. aureus* at recruitment and 211 swab-negative individuals were invited to supply nasal swabs at 2-monthly intervals.^48^ Where co-habiting individuals carried the same *spa*-type, only one individual was considered for inclusion to avoid over-sampling of strains with household transmission. The second was an investigation of nosocomial carriage and transmission at the John Radcliffe Hospital, Oxford between September 2009 and August 2011.^49^ Individuals admitted to three study wards had nasal swabs for *S. aureus* carriage performed on admission and at fortnightly intervals until discharge. 1146 individuals were found to have *S. aureus* carriage on any swab during the study (enrolled from Intensive Treatment Unit (ITU; 729 individuals), Trauma unit (352) or one of two elderly care wards (65)). Carriers with *S. aureus* originally isolated on the first or second day of admission and no overnight stay in the preceding 12 months were classified as community-associated controls (269). Carriers with *S. aureus* originally isolated more than two days after admission and carriers who had been admitted for three or more nights in the preceding 28 days were classified as healthcare-associated controls (335). All carriers with *S. aureus* isolated from a clinical sample in the previous 12 months were excluded. In total 984 asymptomatic carriage controls were successfully sequenced (Figure S2). For carriers with multiple positive samples, the latest sample of the longest carried *spa*-type was selected.

### Epidemiological data

For each episode of SAB, the minimum dataset for inclusion was patient gender and age at the time of infection, date of admission, the number of days between admission and first blood culture from which *S. aureus* was cultured, and the number of days since the most recent discharge from hospital. If available, the clinically-determined focus of infection and vital status 90 days after infection were recorded.

Epidemiological data on episodes of SAB was collected as part of on-going service evaluation studies, as part of multi-centre collaborations with the UK Clinical Infection Research Group (UKCIRG)^6^ and the International *Staphylococcus aureus* Collaboration (ISAC).^7^ Further data, including for carriage controls were obtained from the Infections in Oxfordshire Research Database (IORD) which links information about patient attendances with results from pathology services in an anonymised research database.^50^

Cases were deemed healthcare-associated (HA) if the first blood culture positive for *S. aureus* was collected on the third day or later of a hospital admission (healthcare onset cases), or if the patient had an inpatient admission in the previous 90 days (community onset, healthcare-associated cases).^51^ Cases were deemed community-associated (CA) if the first blood culture positive for *S. aureus* was collected on the first or second calendar day of admission, and there was no inpatient admission in the previous 90 days.

### Microbiological methods

*S. aureus* isolates from blood culture were characterised using standard operating procedures of clinical laboratories at all three centres. Isolates for inclusion were retrieved and frozen in 15% glycerol stock prior to DNA extraction.

Hospital carriage swabs were collected by research nurses using dry cotton-tipped swabs. Community carriage study participants self-collected swabs, returning these by mail as previously described.^48^ All swabs were incubated in 5% saline enrichment broth (Oxoid LTD) overnight at 37°C before subculture onto SaSelect chromogenic agar (Bio-Rad). Plates were examined after 24 hours incubation and potential *S. aureus* colonies confirmed by catalase, DNase and Prolex Staph Xtra Latex kit (Pro-Lab Diagnostics).

Isolates were stored at -80°C in 15% glycerol. Isolates were *spa*-typed as previously reported.^48^

### Whole genome sequencing

For each bacterial culture, a single colony was sub-cultured and DNA was extracted from the sub-cultured plate using a mechanical lysis step (FastPrep; MPBiomedicals, Santa Ana, CA) followed by a commercial kit (QuickGene; Autogen Inc, Holliston, MA), and sequenced at the Wellcome Trust Centre for Human Genetics, Oxford. 600 isolates were sequenced on the Illumina HiSeq 2500 platform (San Diego, California, USA), with paired-end reads 150 base pairs long. The remaining 417 isolates had been sequenced for an earlier on the HiSeq 2000 platform, with paired-end reads of 99 base pairs.

### Variant callin

Following established methods,^17^ we used Velvet^52^ v1.0.18 to assemble reads into contigs *de novo*. Velvet Optimiser v2.1.7 was used to choose the kmer lengths on a per sequence basis. The median kmer length for assembly was 123, however this was affected by sequencing read length, being significantly lower for assemblies based on 99bp reads (median k=69) than those based on 150bp reads (median k=125) (*p*<10^−5^, Wilcoxon rank sum test).

We used BLAST^53^ to find the relevant loci, and defined multilocus sequence type (MLST) using the online database at http://saureus.mlst.net/. Strains that shared six of seven MLST loci were considered to belong to the same Clonal Complex. Antibiotic sensitivity was predicted by interrogating the assemblies for a panel of resistance determinants as previously described.^47^

We used Stampy^54^ v1.0.22 to map reads against a reference genome (MRSA252, Genbank accession number NC_002952).^55^ Repetitive regions, defined by BLAST^53^ comparison of the reference genome against itself, were masked prior to variant calling. Bases were called at each position using previously described quality filters.^39,56,57^ Missing calls were imputed using ClonalFrameML.^58^

### Reconstructing the phylogenetic tree

We constructed a maximum likelihood phylogeny of mapped genomes for visualization using RAxML^59^ assuming a general time reversible (GTR) model, and fine-tuned the estimates of branch lengths using ClonalFrameML.^58^

### Kmer counting

We used a kmer-based approach to capture non-SNP variation.^60^ Using the *de novo* assembled genomes, all unique 31 base haplotypes were counted using dsk.^61^ If a kmer was found in the assembly it was counted present for that genome, otherwise it was treated as absent. This produced a set of 23,860,793 variably present kmers, with the presence or absence of each determined per isolate. We identified a median of 2,760,000 kmers per isolate, including variably present kmers and kmers common to all genomes (IQR 2,725,000 - 2,806,000). The number of kmers found per isolate did not differ significantly with sequencing platform (*p*=0.4, Wilcoxon rank sum test). From a smaller set of 1610 isolates sequenced with 150bp reads, we identified 22,284,204 variably present kmers.

### Calculating heritability

We used the Genome-wide Efficient Mixed Model Association tool (GEMMA^62^) to fit a linear mixed model for association between a single phenotype (bacteraemia vs asymptomatic nasal carriage, [encoded as 1 and 0, retrospectively]). We calculated the relatedness matrix from biallelic SNP and kmer presence for tests of each allele type. We used GEMMA to estimate the proportion of variance in phenotypes explained by genotypic diversity (i.e. heritability).

### Genome wide association testing of SNPs and Kmers

We performed association testing using an R package bacterialGWAS (https://github.com/jessiewu/bacterialGWAS), which implements a published method for locus testing in bacterial GWAS.^63^ The association between each SNP and kmer with the phenotype was tested controlling for population structure and genetic background using the linear mixed model (LMM) implemented in GEMMA. ^62^ We included healthcare or community origin of case/control status as a fixed covariate in the model when testing for associations with the carriage-vs-bacteraemia phenotype. The parameters of the linear mixed model were estimated by maximum likelihood and a likelihood ratio test was performed against the null hypothesis (that each locus has no effect) using the software GEMMA, ^62^ using a minor allele frequency of 0 to include all SNPs. GEMMA was modified to output the ML log-likelihood under the null and alternative hypothesis and –log_10_ *p* values were calculated using R scripts in the bacterialGWAS package.

### Testing for lineage effects

We tested for associations between lineage and phenotype using principal components (PCs) in the R package *bugwas* (available at https://github.com/sgearle/bugwas), which implements a published method for lineage testing in bacterial GWAS. ^63^ PCs were computed based on biallelic SNPs using the R function prcomp. To test the null hypothesis of no background effect of each principal component, we used a Wald test^64^ against a *χ*^2^ distribution with one degree of freedom to obtain a *p* value.

### Kmer mapping and sequence alignment

We used Bowtie^65^ to align all 31bp kmers from short-read sequences to the reference genome (MRSA252^55^). For all 31bp kmers significantly associated with case-controls status, the likely origin of the kmer was additionally determined by nucleotide sequence BLAST^53^ of the kmers against a database of all *S. aureus* sequences in Genbank. We used BLAST^53^ to identify the best match for coding sequences of interest in the *de novo* assembly, and used Jalview^66^ to and visualize the assembled sequences.

### Multiple testing correction

Multiple testing was accounted for by applying a Bonferroni correction;^67^ the individual locus effect of a variant (PC, kmer or SNP) was considered significant if its *P* value was smaller than *α*/*n*_p_, where we took *α* = 0.05 to be the genome-wide false-positive rate and *n*_p_ to be the number of PCs, kmer phylopatterns or SNP phylopatterns. We defined each phylopattern to be a unique partition of individuals by the alleles at that kmer or SNP.

The Bonferroni correction represents a conservative approach to controlling for type 1 error. The harmonic mean *p*-value (HMP) has recently been developed as a method to combine alternative hypotheses against the null hypothesis, without sacrificing power, even when the tests are not independent.^68^ The HMP was calculated for coding regions using the R package “harmonicmeanp” v3.0 (https://CRAN.R-project.org/package/harmonicmeanp). The HMP across a region was then adjusted for the proportion of kmers mapping to that region:

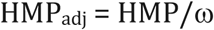

(where ω = proportion of kmers mapping to that region, compared to the total number of kmers mapping to coding regions). The adjusted HMP was compared directly to the significance threshold *α*_L_, being a nominal threshold *α* (0.05) adjusted for the number of unique *p*-values being tested.^68^

## Results

Sequences from 2001 *S. aureus* isolates (1,017 cases of bacteraemia and 984 asymptomatic nasal carriage controls) were analysed (Table 1). Cases were marginally more likely to be healthcare-associated (38% vs33.5%, *p*=0.04). Consistent with established risk factors for SAB,^2^ cases were significantly older (median age 68 years vs 59 years, *p*<10^−5^) and more likely to be male (68.4% vs 51.9%, *p*<10^−5^). Cases had a higher proportion of MRSA than controls (13.6% vs 5.5%, *p*<10^−5^), including when comparing HA cases (63/386 (15.3%)) with HA controls (35/330 (10.6%), *p*=0.04 (χ^2^ test)). Thus, even in individuals exposed to the healthcare environment, MRSA was found more often in bacteraemia than carriage. Reported focus of infection in cases of SAB showed soft tissue and vascular catheter infections to be the most commonly identified foci (Table S1). Mortality by 30 days was 26.5% (Table S2). These observations are consistent with previously reported UK cohort studies of SAB.^6,7,8^

**Table 1:** Cases and controls included in study. HA includes both hospital-onset disease and community-onset, healthcare acquired disease.

|  | ***S. aureus***<br>**bacteraemia (cases)** | ***S. aureus* nasal**<br>**carriage (controls)** |  |
| --- | --- | --- | --- |
| Number of sequences | 1017 | 984 |  |
| Community-associated (CA) | 631 (62.0%) | 654 (66.5%) |  |
| Healthcare-associated (HA) | 386 (38.0%) | 330 (33.5%) | $p = 0.04$ ($\chi^2$ test) |
| Age (median (IQR)) | 68 (53-79) | 59 (38-76) | $p < 10^{-5}$ (Mann-Whitney U test) |
| Male sex (n (%)) | 659 (68.4%) | 511 (51.9%) | $p < 10^{-5}$ ($\chi^2$ test) |
| MRSA (n (%)) | 138 (13.6%) | 54 (5.5%) | $p < 10^{-5}$ ($\chi^2$ test) |

### *S. aureus* lineages do not differ strongly in their propensity to cause bacteraemia

A phylogeny of 2001 cases and controls demonstrated that a broad diversity of *S. aureus* lineages among our cases (Figure 1), with representatives from all clonal complexes (CC) as defined in the MLST scheme.^69^ Cases and controls did not obviously cluster in the tree. Two lineages dominated MRSA isolates – ST-22 (within CC-22) and ST-36 (within CC-30) – consistent with the epidemiology of MRSA in Oxfordshire and throughout the UK.^34,70,71,72^ HA cases and controls were distributed throughout the tree, and did not strongly cluster within the population.

**Figure 1:**
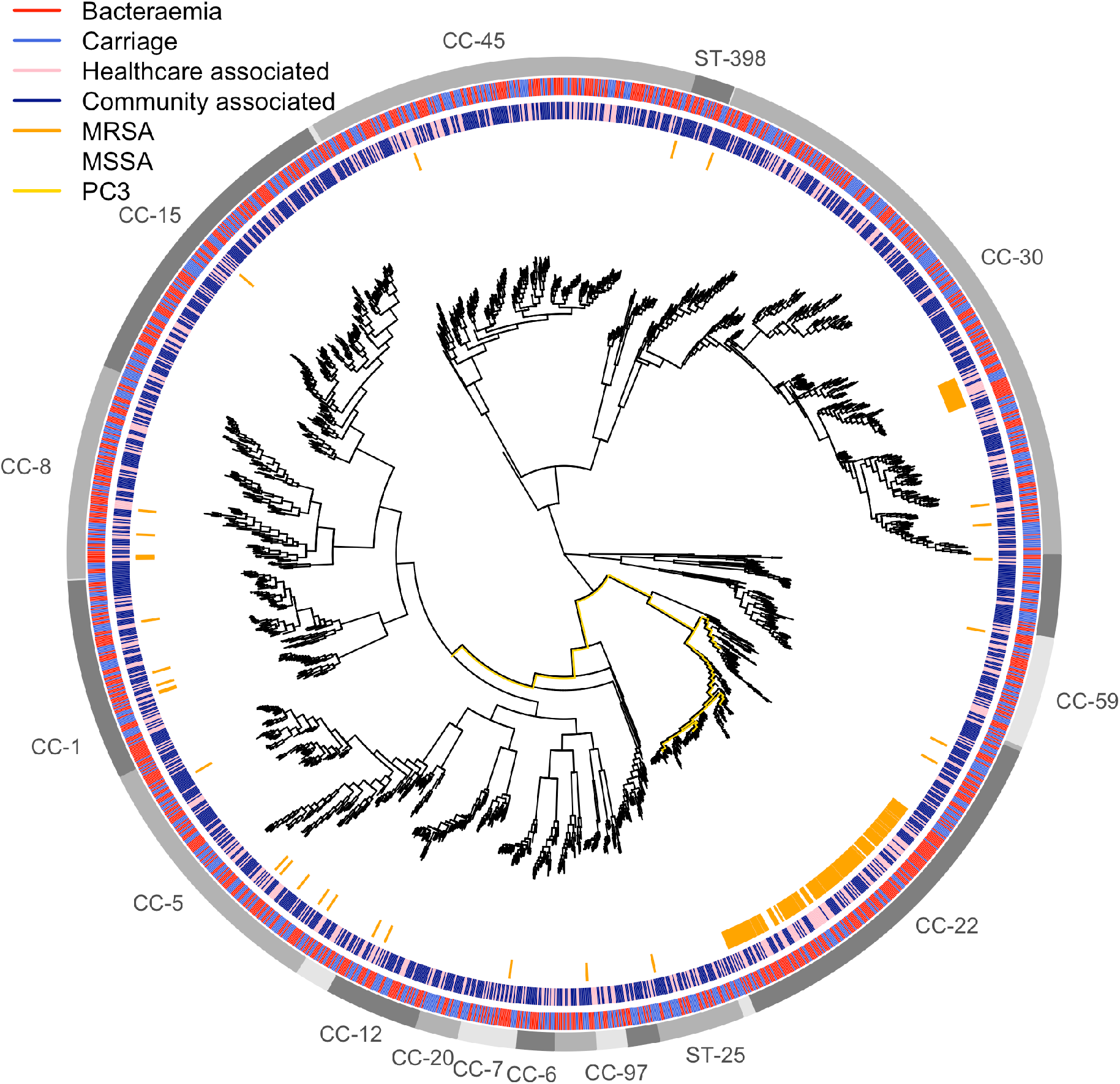
Maximum likelihood phylogeny of 2001 isolates from bacteraemia and carriage. Branch lengths have been square-root transformed to better discriminate closely related lineages. The outer ring indicates clusters with a shared lineage; lineages with more than 20 isolates are named by the clonal complex (or ST if only a single ST was in the cluster). The second outermost ring indicates isolate source (blue carriage, red bacteraemia). The third outermost ring indicates whether each isolate was community (dark blue) or hospital (pink) associated. The inner ring indicates isolates were MRSA (orange) or MSSA (white). The branches corresponding with the most significant principal component of variance (PC3, *p*=0.02, Wald test) with respect to carriage-vs-bacteraemia are highlighted in yellow.

Formal testing for lineage effects with *bugwas*^63^ supported the absence of strong lineage effects. The third principal component (PC3), which identified the MRSA clade within the CC-22 lineage, was most strongly associated with bacteraemia-vs-carriage (*p=*0.02), but this was not statistically significant after adjusting for multiple testing (Figure 1, Figure S3). The overall sample heritability was predicted to be low (2.1%, 95% CI 0.0-5.3%). This comprehensive survey of SAB and carriage indicates that lineages of *S. aureus* do not differ substantially in their intrinsic propensity to cause bacteraemia.

### Antimicrobial resistance determinants are associated with *S. aureus* bacteraemia

To investigate associations between genomic content or sequence variation and the carriage-vs-bacteraemia phenotype we tested SNP associations and also used a kmer approach,^60^ identifying 31bp DNA sequences (kmers) in the assembled genome and testing for association between their presence and bacteraemia. This approach allowed us to detect variation in the accessory genome, and variants such as small insertions or deletions that are not well captured by mapping SNPs.

Testing all identified SNPs for association with case/control status in 2001 isolates did not identify any statistically significant associations between individual SNPs and bacteraemia at the genome wide level when controlling for population structure (Figure S4). However, the SNP coming closest to a statistically significant association (*p=*10^−5.6^) was an A to T mutation at position 1497290 in the MRSA252 reference genome. This SNP encodes a phenylalanine to tyrosine substitution at codon position 99 in dihydrofolate reductase (*dfrB*); this F99Y mutation confers trimethoprim resistance.^47^ It was relatively rare, being found in 41 cases and 5 controls, and correlated with resistance to other antibiotics: 36/46 (78%) of isolates with this variant were also methicillin resistant. This variant was found most commonly in isolates from CC-22 (63%) and ST-36 (22%).

When testing kmers found in all 2001 isolates, we found 1214 kmers significantly associated with carriage. These kmers mapped to multiple sites across the genome, most of which were repeat regions, including 16S rDNA and transposon insertion sequences (Figure S5A). However, the presence of these kmers was strongly affected by the Illumina sequencing read length, being found in isolates sequenced using 150bp but not 99bp reads (Figure S5B). *De novo* assembly was repeated using a constrained kmer length in assembly (up to 79bp) to control for the variation in read length, and when these new assemblies were used, 76% (924/1214) of the previously identified 31bp kmers were no longer significantly associated with the phenotype (Figure S5c). We concluded that varying length of sequencing was a major source of confounding in kmer-based associated estimates.

To avoid false positive results, we therefore restricted the investigation of kmers associated with carriage-vs-bacteraemia to isolates sequenced with 150bp reads (Table 2). This reduced set of cases showed similar epidemiological characteristics to the larger group (Table 1).

**Table 2:** Cases and controls sequenced at 150 bp read-length and included in the kmer study. HA includes hospital-onset disease and community-onset, healthcare-acquired disease.

|  | ***S. aureus***<br>**bacteraemia (cases)** | ***S. aureus* nasal**<br>**carriage (controls)** |  |
| --- | --- | --- | --- |
| Number of sequences | 626 | 984 |  |
| Community-associated (CA) | 410 (65.5%) | 654 (66.5%) |  |
| Healthcare-associated (HA) | 216 (34.5%) | 330 (33.5%) | $p = 0.7$ ($\chi^2$ test) |
| Age (median (IQR)) | 68 (53-79) | 59 (38-76) | $p < 10^{-5}$ (Mann-Whitney U test) |
| Male sex (n (%)) | 399 (63.7%) | 511 (51.9%) | $p < 10^{-5}$ ($\chi^2$ test) |
| MRSA (n (%)) | 78 (12.4%) | 54 (5.5%) | $p < 10^{-5}$ ($\chi^2$ test) |

Kmers tagging antimicrobial resistance (AMR) conferring mutations were significantly associated with bloodstream infection. In total, we identified 22,284,204 kmers, occurring in 930,702 unique patterns across 1610 isolates with 150bp reads. Twenty-three kmers, occurring in two phylopatterns, were significantly associated with SAB (Figure 2). These kmers, mapping to a 52bp region in *dfrB*, exhibited 11.2-11.6-fold increased odds of being found in a disease-causing, rather than carried, *S. aureus*. This association remained significant after controlling for population structure (*p=*10^−8.9^-10^−9.3^). When mapped, these kmers centred on MRSA252 position 1497290, where three known single nucleotide variants are capable of conferring trimethoprim resistance, including the F99Y variant identified by our SNP GWAS. Like the trimethoprim resistance conferring SNP, these kmers were found in low frequencies (35/626 cases and 5/984 controls).

**Figure 2:**
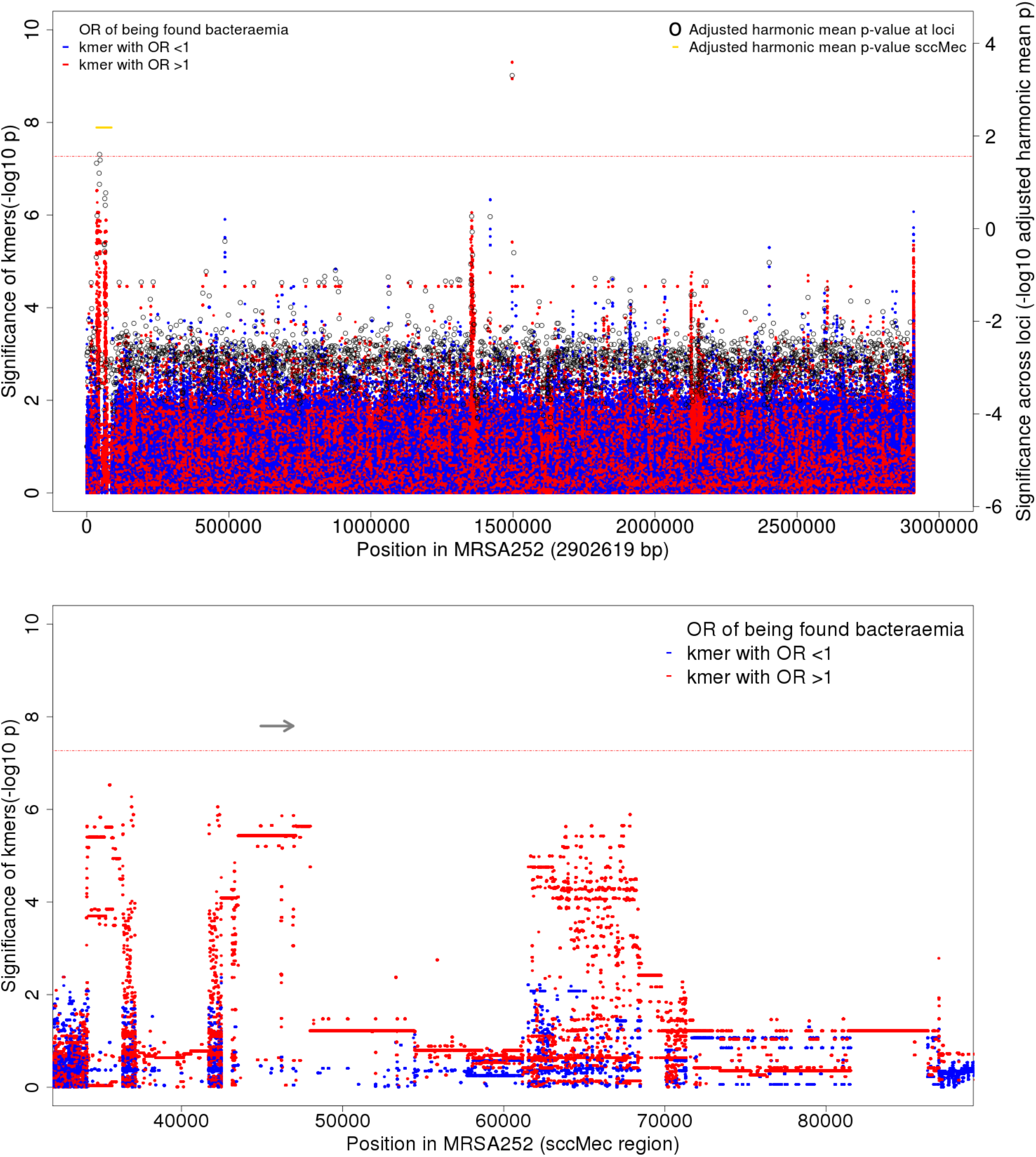
Association of kmers with *S. aureus* bacteraemia-vs-carriage, controlling for population structure and HA or CA origin. (A)Manhattan plot showing significance of association (-log10 *p*-value, left axis) for individual kmers (red, kmers with OR >1 of being found in bacteraemia-vs-carriage; blue, kmers with OR <1 of being found in bacteraemia-vs-carriage). Unmapped kmers are plotted at the end of the genome. Pooled evidence (adjusted harmonic mean *p*-value, right axis) across each CDS is shown by black open circles. Pooled evidence across SCCmec is shown in gold. A threshold of significance is plotted in a red horizontal line. For kmers this a Bonferroni-corrected threshold of significance, adjusted for the number of individual kmer patterns (10^−7.2^, left axis). For evidence across loci, the same family wide error rate of 0.05 (10^−1.6^, right axis) was applied by adjusting the p-values for the numbers of variants tested. (B) Kmers mapping to the region of staphylococcal chromosome cassette sccMec are shown in greater detail. The coding sequence of *mecA* (SAR0039) is marked by a horizontal grey arrow.

No further individual kmers met the threshold for significance, but there were distinctive peaks enriched for small *p*-values in the Manhattan plot (Figure 2). We calculated harmonic mean *p*-values (HMP) to perform aggregate kmer-based tests of association across coding regions of the genome. The HMP can improve power by combining information across variants and reducing the effective number of tests, thereby attracting a less stringent significance threshold. At the whole-genome level, the pooled evidence for association between coding sequence variation and bacteraemia was considerable (HMP=10^−3.3^). The evidence for kmers associated with bacteraemia, when pooled, was significant in several loci (Figure 2), including the *dfrB* locus, SAR1439 (HMP=10^−6.9^, HMP_adj_=10^−3.3^). The presence of kmers mapping across the SCCmec region was also significantly associated with bacteraemia (HMP=10^−4.4^, HMP_adj_=10^−2.2^). The strongest evidence within SCCmec was for *mecA* (SAR0039, HMP=10^−5.3^, HMP_adj_=0.02), which encodes the low affinity PBP2a that confers methicillin resistance.

When this region was examined in closer detail, high-risk kmers covered the entirety of the SAR0039 locus, encoding PBP2a, a PBP with low affinity for beta-lactams (Figure 2B). There are no alternate low-risk kmers in this region, suggesting the presence of this gene, rather than variation within it, is associated with bacteraemia. Thus, genomic sequences associated with AMR to both trimethoprim and beta-lactams, but not other antimicrobial classes, were significantly associated with bacteraemia.

### Genomic signals of healthcare-associated carriage include antimicrobial resistance factors, as well as variation in a virulence determinant, *prsA*

Regulatory gene changes have been associated with persistent bacteraemia and increased mortality, and it has been hypothesised these changes either convey or accompany relative survival advantages in healthcare environments.^31,32^ We compared the genomic factors associated with healthcare environments by conducting a GWAS for HA-vs-CA among carriers (330 HA, 654 CA). We focused only on carriage isolates because we had more extensive data about hospital admissions in this group, and epidemiological data to further demonstrate that isolates from CA-carriage truly reflected community origin. The MRSA CC-22 lineage showed the strongest association with HA-vs-CA carriage (*p*=0.06, Figure S7), but this lineage effect was even less significant than for bacteraemia-vs-carriage (*p*=0.02, Figure S3), suggesting that no lineages were strongly associated with healthcare acquisition of *S. aureus* carriage.

In total 124 SNPs were associated with HA-vs-CA carriage after controlling for population structure, and adjusting for the 77,597 SNP phylopatterns in the population (Figure S8, Table S3). The most significant were non-coding (intergenic) variants in a region encoding tRNAgly at 2034022-2034039. However, basecalls at these sites were frequently imputed (61.5%). Excluding isolates with imputed calls at these sites reduced the unadjusted OR for finding these SNPs in HA-vs-CA carriage from 2.1 to 0.87, suggesting that the observed association was a product of SNP imputation. The most significant coding variants included a G to A substitution at 2417648 in MRSA242 (*p*=10^−7.2^), encoding a proline to leucine substitution at 707 in SAR2345, an AcrB/AcrD/AcrF family protein, which is a multidrug efflux system subunit. They also included a C to T substitution at 7255 in MRSA252 (*p*=10^−7.2^), which encodes L84S in *gyrA* and confers quinolone resistance.^47^ A variant at these positions was called in all 984 isolates, so no calls were imputed. A significant association was also seen with a band of 91 low frequency SNPs, with shared minor alleles co-inherited predominantly in the MRSA sub-clade of CC-22 (Figure S9). The *dfrB* mutation seen associated with bacteraemia was not significantly associated with HA-vs-CA carriage (*p*=0.4).

188 31bp kmers in 59 unique patterns were significantly associated with HA carriage (Figure 3A, Table S4); 123 (65.4%) kmers comprising 52 phylopatterns mapped to a single gene – *prsA* – and their absence was associated with HA carriage (*p=*10^−7.2^-10^−19.4^)(Figure 3B). A small peak of 11 significant kmers mapped to a hypothetical protein SAR0061, covering to a 41bp region from 67,816 (*p*=10^−7.3^). Three significant kmers mapped to a putative transcriptional regulator SAR2394, covering a 33bp region at 2,465,937(*p*=10^−7.2^). Of the remaining significant kmers, 26/188 (13.8%) did not map to the reference genome, and 25/188 (13.3%) mapped to non-coding regions at 1.3MB, 2.7MB and 2.8MB.

**Figure 3:**
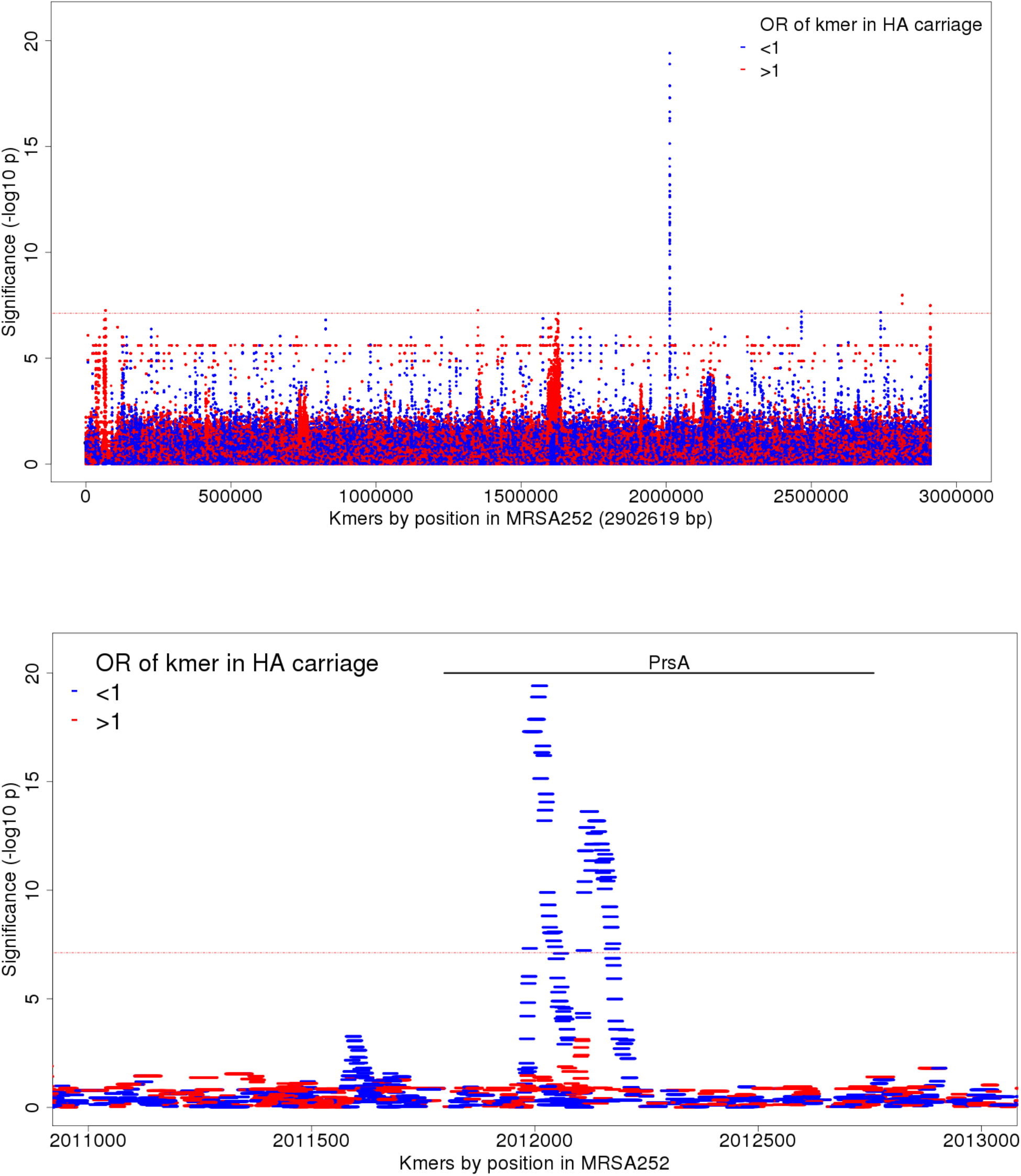
Association of kmers with HA-vs-CA carriage, controlling for population structure. (A)Manhattan plot showing significance of association (-log10 *p*-value, left axis) for individual kmers (red, kmers found more often in HA carriage; blue, kmers found more often in CA carriage). The Bonferroni significance threshold, adjusting for the number of kmer phylopatterns, was 10^−7.1^ (red horizontal line). (B) The region of *prsA* is shown in greater detail. Kmers showing significant association with HA-vs-CA carriage cover the region 2011973-2012202 in the reference, which corresponds to bases 177-394 in prsA

While CA carriage isolates showed conservation of the *prsA* sequence across the population, multiple patterns of variation arising in different lineages were seen in HA carriage isolates (Figure S10), consisting of both SNPs and deletions (Figure S11). When a conditional SNP GWAS was performed including the presence or absence of any of the kmers most strongly associated with CA-vs-HA carriage as a covariate in the model, no SNPs reached the threshold for significance, suggesting the signal of association accompanying these SNPs was better explained by *prsA* variation. Variation in *prsA* was common in the MRSA sub-clade of CC-22 but was not limited to this lineage. PrsA is a surface bound foldase protein, responsible for post-translational processing of virulence factors, including proteases and cell surface proteins^80,81^ which has also been shown to modulate susceptibility for both beta-lactam and glycopeptide antibiotics.^78,79^ No peak of significant kmers occurred at the *prsA* locus in the carriage-vs-bacteraemia GWAS (Figure 2A), suggesting that variation at this locus is specifically associated with carriage arising from the hospital environment, but not with bacteraemia.

## Discussion

These findings reflect analysis of a large collection, representing the populations of *S. aureus* circulating in community and hospital settings, including the whole genomic content of the population, with control of population structure. In doing so, we address the conflicting results from smaller case-control studies which have found evidence for^26,29^ and against^18,24^ differing invasiveness between *S. aureus* lineages. Broadly, lineages did not differ in the frequency with which they caused bacteraemia, compared to their frequency in carriage. However, across lineages we found evidence that genetic variants underlying AMR were associated with increased odds of bacteraemia versus carriage. These included some determinants of methicillin resistance, contrasting with previous research indicating that methicillin resistance incurs a fitness cost for *S. aureus*, reducing its pathogenicity.^82,83^

The most obvious possible explanation for association between methicillin resistance and bacteraemia would be survival benefit in the presence of beta-lactam antibiotics. However more than half of our MRSA bacteraemia cases were community-associated, being first detected prior to or within 48 hours of hospital admission. While we do not have data about pre-hospital antibiotic treatment, it is unlikely that the majority of patients with CA bacteraemia were on beta-lactam antibiotics at the time that bacteraemia developed. It is also possible that other between-group differences, such co-morbid illnesses, may also account for the different prevalence of MRSA observed between patients with bacteraemia and asymptomatic carriers, and we have not been able to measure patient factors confounding between MRSA carriage and invasive infection.

However, there is *in vitro* evidence that *mecA* – the primary determinant of methicillin resistance – modifies bacterial virulence independently of antibiotic selection pressure, through modulation of Agr-mediated toxin expression.^33^ The association observed here concords with other evidence implicating the altered PBP encoded by *mecA* in persistent and complicated bacteraemia. Analysis of SAB over 21 years in the USA showed that within CC-8, the MRSA lineage USA300 was associated with higher rates of metastatic infection compared to the rest of the *spa*-t008 complex after adjusting for patient and clinical variables.^73^ *S. aureus* strains which are phenotypically methicillin susceptible but contain the *mecA* gene have been reported to have a higher risk of persistent bacteraemia, even when treated with vancomycin.^74^ AMR elements have been implicated as virulence determinants in another Gram positive species; *Streptococcus pneumoniae*, where a PBP variant associated with penicillin tolerance (though not resistance) was associated with meningitis among isolates found in invasive pneumococcal disease.^75^

Our finding of trimethoprim resistance associated with bacteraemia is not easily explained through direct antibiotic selection since trimethoprim containing antibiotics (e.g. co-trimoxazole) are not advised in therapeutic guidelines for treatment of skin and soft tissue infections in the United Kingdom, where CA-MRSA rates are low. It is plausible that patients may receive trimethoprim for urinary tract infections in the community, and this could reflect an impact on commensal flora, enabling invasion by trimethoprim-resistant *S. aureus* strains. It is also possible that mutations in *dfrB*, a metabolic gene important in bacterial DNA synthesis, affect bacterial persistence: trimethoprim resistant *S. aureus* have shown variable growth rate and survival under environmental stress according to the mechanism of resistance.^77^

Overall, we found MRSA was more prevalent in bacteraemia than carriage in samples, when sampling the population representatively. In the UK at this time MRSA was almost exclusively healthcare associated. Consequently, one potential explanation for this finding is unmeasured hospital exposure among CA bacteraemia cases for which hospital exposure data was only available for the preceding 12 weeks. However even restricting to HA associated cases and controls MRSA was associated with bacteraemia. Furthermore, by controlling for population structure, we can be confident the association demonstrated between *mecA* and bacteraemia is not simply a reflection of hospital adapted lineages, and that such lineage effects could have been detected using the GWAS methods employed here. In fact, while variation within *prsA* was strongly associated with HA carriage, variation in this gene was not associated with carriage-vs-bacteraemia. Likewise, a quinolone resistance mutation in *gyrA* was associated with HA carriage but not with bacteraemia. These observations suggest that distinct bacterial factors favour hospital adaptation or transmission compared with those favouring bloodstream infection.

The surface bound foldase protein PrsA has been implicated as a secondary resistance factor for both beta-lactam and glycopeptide antibiotics.^78,79^ *In vitro* assays have shown that *S. aureus* can survive despite disruption to *prsA*, but strains with *prsA* disruption are more susceptible to oxacillin.^78^ Regulating expression of a methicillin-resistant phenotype independently of *mecA*, PrsA reduces the membrane quantity of PBP2a without altered transcription of *mecA*.^79^ Additionally, PrsA plays an important role in the post-translational processing of virulence factors, including proteases and cell surface proteins,^80,81^ and while the secretion of some virulence factors is decreased when *prsA* is deleted,^80^ PrsA-deficient bacteria have enhanced aggregation and adherence,^81^ changes which might favour survival or transmission in the hospital environment.

Our study demonstrates some constraints and pitfalls for bacterial GWAS. Firstly, sequencing read length was a strong source of confounding in our data which, without adequate control, produced false positive results. This is an ongoing challenge for studies pooling existing sequencing data where it is not possible to randomize cases and controls across sequencing batches. We dealt with this using the conservative option of excluding 391 cases from kmer analysis, representing a substantial sacrifice of power. A further limitation was that population structure appeared to be incompletely controlled for low frequency variants,^84^ as exhibited by a set of 91 SNPs in strong linkage disequilibrium (LD) associated with HA-vs-CA carriage. Linear Mixed Models are able to control for lineages, and cryptic population structure through the use of a relatedness matrix, but they make the assumption that closely related isolates are unlikely to have large differences in phenotype. This assumption means that they can incompletely account for lineage effects arising from closely related strains which vary significantly in frequency between phenotypes.^84^ In our study, the SNPs in LD were found in the predominantly HA-MRSA isolates within CC-22. This may represent either a true association with that lineage, or co-carriage with another genomic element in that lineage. In this case, *prsA* variation was common in CC-22, but not detected in the SNP based study (as the variation was a deletion rather than a nucleotide substitution), and the signal of association accompanying these SNPs was better explained by *prsA* variation. Overall, the kmer-based methods were more fruitful in our study, both in their ability to detect non-SNP based variation (such as deletions in *prsA*, and the presence of *mecA*), and retaining the ability to identify significant SNPs (including a *dfrB* variant).

In previous studies we have identified within-host adaptation of *S. aureus* genes associated with development of invasive disease from a colonising isolate – particularly involving the *agr* locus, and the cell wall proteins under regulatory control of Agr and Rsp.^43^ Such variation was not associated with bacteraemia in this population-based study, perhaps because it provides only a short-term advantage to the bacteria, and in the longer term is detrimental, by adversely affecting transmission. In contrast, variation which confers AMR is likely to confer a bacterial survival advantage in carriage in the face of antibiotic selection pressure in the healthcare environment, as well as in the bloodstream, allowing these variants to survive in the population.

## Conclusions

In a study of over 2000 isolates from *S. aureus* bacteraemia and nasal carriage in the UK, we found strong evidence that all *S. aureus* lineages are equally capable of causing bloodstream infection, and of being carried in the healthcare environment.

We found that genomic variation in the foldase protein *prsA* was a novel genomic marker of healthcare adaptation in *S. aureus*. This predictor of healthcare-associated carriage was not associated with bacteraemia, while AMR determinants were associated with both bacteraemia and hospital-associated carriage, raising the suggestion that in addition to enabling survival in healthcare environments, AMR functions as a virulence factor, promoting invasive disease.

Given studies demonstrating a direct effect of *mecA* on toxin expression^33^ and reduced toxicity enhancing bloodstream survival,^42^ we hypothesise that lowered expression of toxic virulence factors seen in MRSA may be one method by which *S. aureus* gains a short-term survival advantage and causes bloodstream infection.

## Data Availability

Sequenced bacterial isolates submitted to Short Read Archive accession number PRJNA690682

## Abbreviation

Agr: Accessory gene regulator
AMR: Antimicrobial resistance
CA: Community associated
CC: clonal complex
HA: Healthcare associated
HMP: Harmonic mean *p*-value
MLST: Multi locus sequence type
MRSA: Methicillin-resistant *S. aureus*
LD: linkage disequilibrium
OR: Odds ratio
PBP: Penicillin binding proteins
PVL: Panton Valentine leucocidin
Rsp: Repressor surface protein
SAB: *Staphylococcus aureus* bacteraemia
*S. aureus*: *Staphylococcus aureus*
WGS: Whole genome sequencing

## Declarations

### Ethics approval and consent to participate

The study of community *S. aureus* carriage was approved by Oxfordshire Research Ethics Committee B (08/H0605/102). Ethical approval for linkage to patient data without individual patient consent in Oxford was obtained from the South Central Ethics Committee (14/SC/1069) and the Confidentiality Group [ECC5-017(A)/2009]. Data about *S. aureus* bacteraemia in Oxfordshire, Brighton and Plymouth were collected for evaluations of clinical service provision. Under UK National Research Ethics Service guidance at the time this data collection constituted a service evaluation involving routinely available, non-identifying patient data and therefore not requiring research ethics committee review.

### Consent for publication

Not applicable

### Availability of data and materials

sequenced bacterial isolates deposited in Short Read Archive accession number PRJNA690682

### Authors’ contributions

Study was designed by BCY, CHW, JC, SE DCW, TEAP, ASW, MJL, DJW. Samples and epidemiological information collected by BCY, JRP, JCG, SO, HG, RF, AV, RM, KK, TPQ, RT, MS, MJL, sequencing performed by KC, LD, EL, BCY. Analysis of data by BCY, CHW, JC, SE, ASW, MJL, DJW. Discussion of results by BCY, CHW, DWC, TEAP, ASW, MJL, DJW. Manuscript prepared by BCY and DJW.

## Acknowledgements

We thank the International *Staphylococcus aureus* Consortium and the United Kingdom Clinical Infection Research Group for sharing data and bacterial strains from bloodstream infection.

---

**Figure S1:**
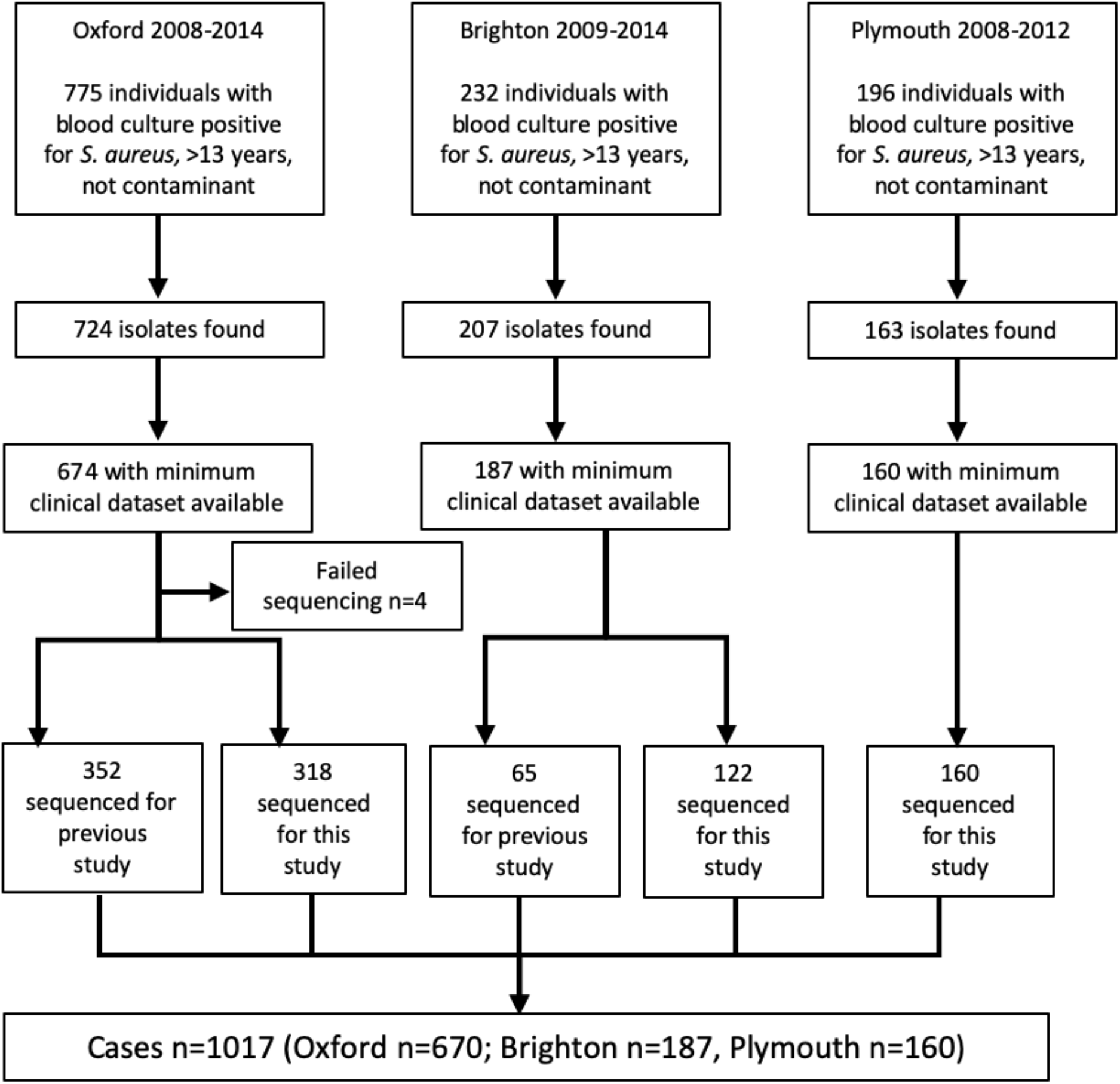
Flow diagram of cases screened for inclusion in the study.

**Figure S2:**
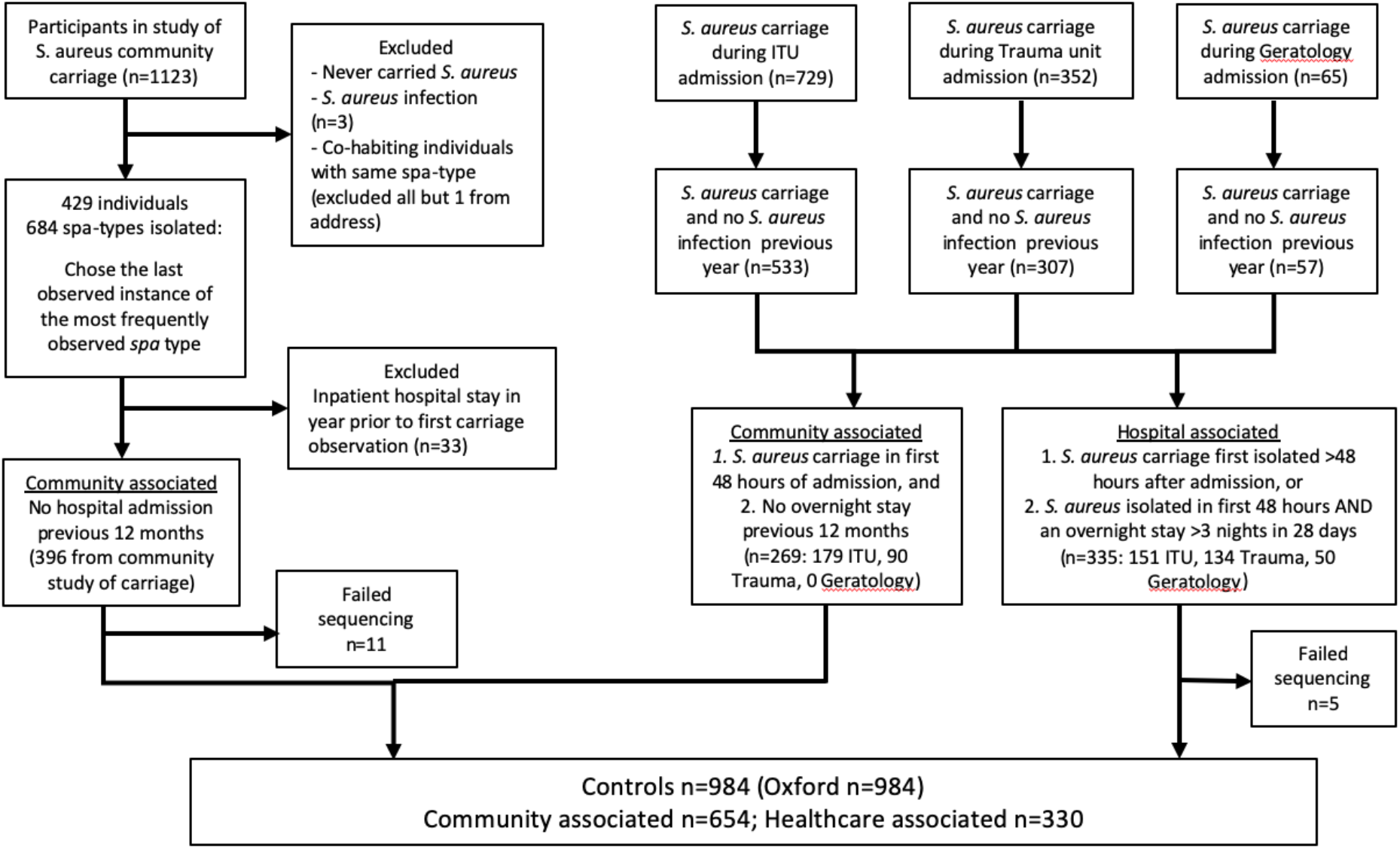
Flow diagram of controls screened for inclusion in the study.

**Figure S3:**
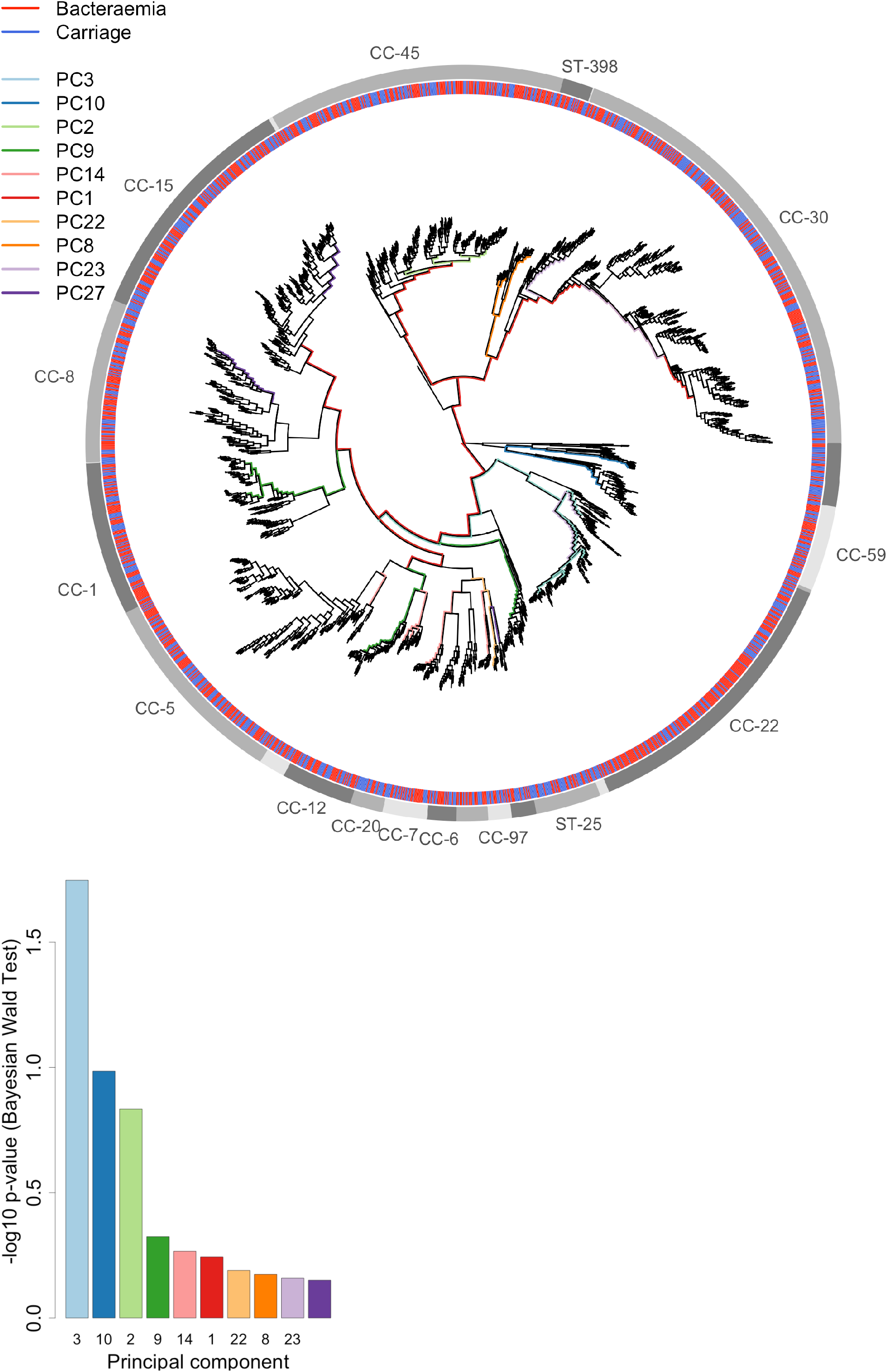
Association of Principal Components with carriage-vs-bacteraemia. **(A)** The branches corresponding to the 10 lineages most significantly associated with carriage-vs-bacteraemia are coloured on a maximum likelihood phylogeny of the study isolates. Branch lengths have been square-root transformed to discriminate closely related lineages. Clonal complexes are denoted in the outermost ring. The second outermost ring indicates isolate source (blue carriage, red bacteraemia) **(B)** Significance (negative log_10_ *p*-values) of the 10 most significantly associated PCs. A Bonferroni corrected threshold for significance is 10^−4.6^.

**Figure S4:**
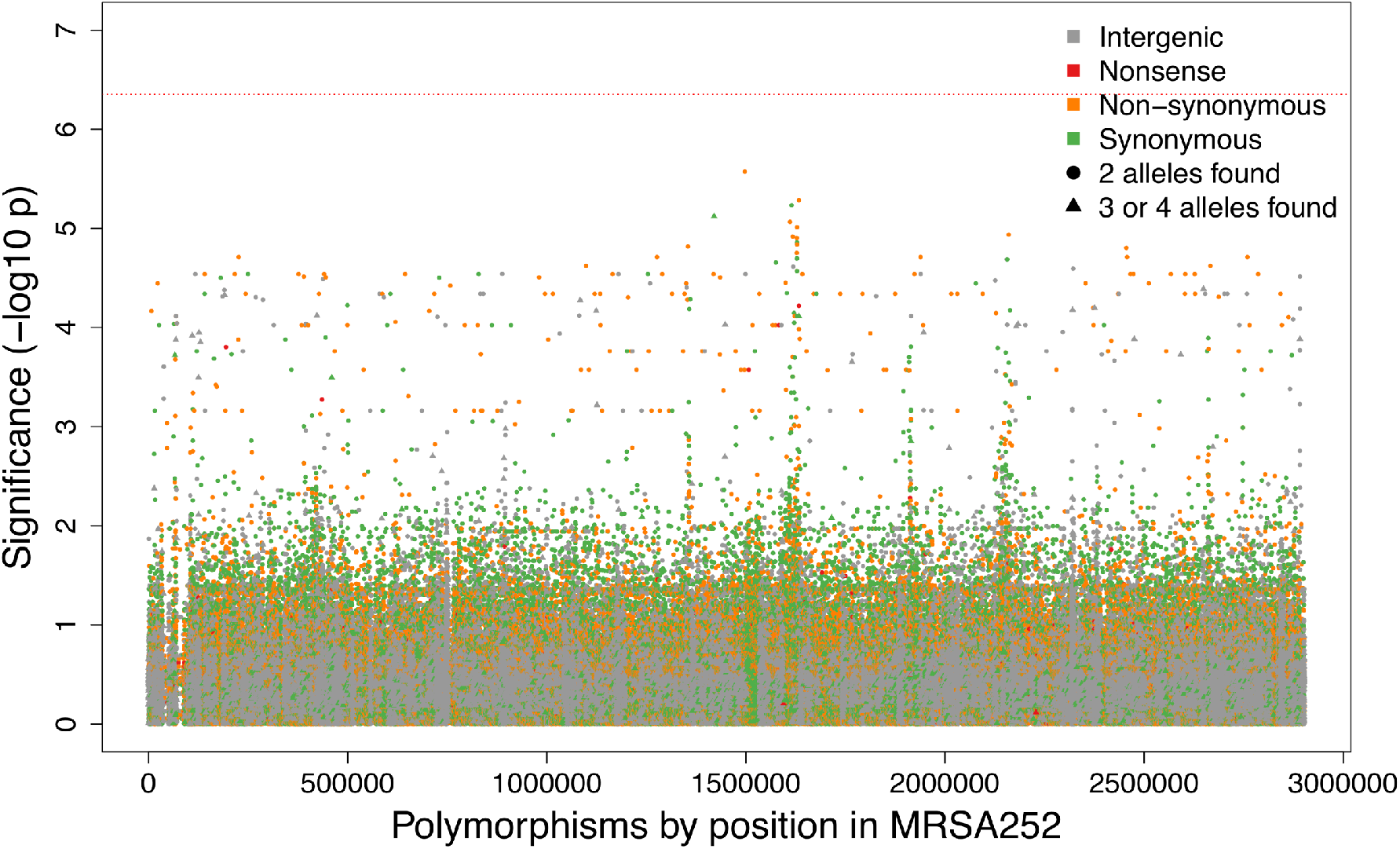
Manhattan plot of SNP associations with carriage-vs-bacteraemia, controlling for population structure and HA or CA origin. The significance of each SNP (–log_10_ *P* value) is plotted according to location on the 2.9MB MRSA252 reference genome, with control for population structure and CA-vs-HA origin. SNPs are coloured according to their predicted effect on protein, and shaped according to the number of alleles found at that site in the study set. A Bonferroni-corrected significance threshold based on the number of phylopatterns (n=112, 292) is plotted in red (10^−6.4^), assuming a family-wise error rate of 5%.

**Figure S5:**
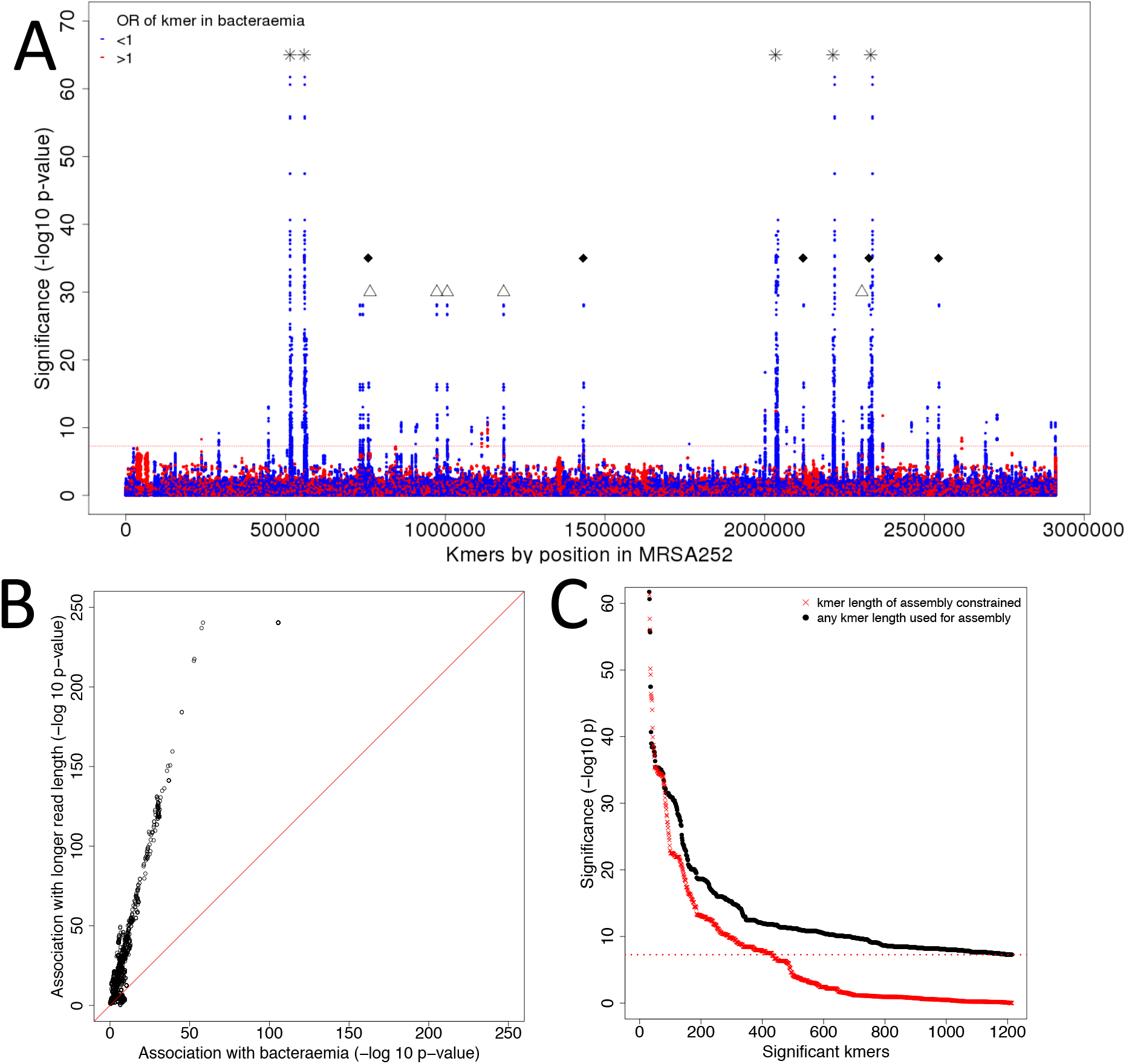
Confounding of kmer study by sequencing read length. **(A)** Manhattan plot of kmers associated with phenotype in study of 2001 isolates. Kmers are mapped to reference genome and plotted against statistical significance of association with case-control-status. Kmers found more frequently in carriage are mapped in blue, kmers found more frequently in bacteraemia are mapped in red. A Bonferroni corrected threshold for significance is plotted in red (10^−7.3^). The positions of several repeat regions are marked: 16S rDNA (*), ISS (Δ) and ISSSau3 (♦). **(B)**The kmers most associated with bacteraemia-vs-carriage were confounded with sequence length. We plotted significance of association with both bacteraemia-vs-carriage (X axis) and sequencing read length (Y axis), using a χ^2^ test, without correction for population structure. **(C)** Kmer detection and association testing with GEMMA was repeated, this time on new assemblies, built using Velvet with a maximum kmer length of 79bp. The signal of the most significant kmers based on the original assemblies (black) dropped to below statistical significance when the assemblies were built with a constrained kmer length (red).

**Figure S6.**
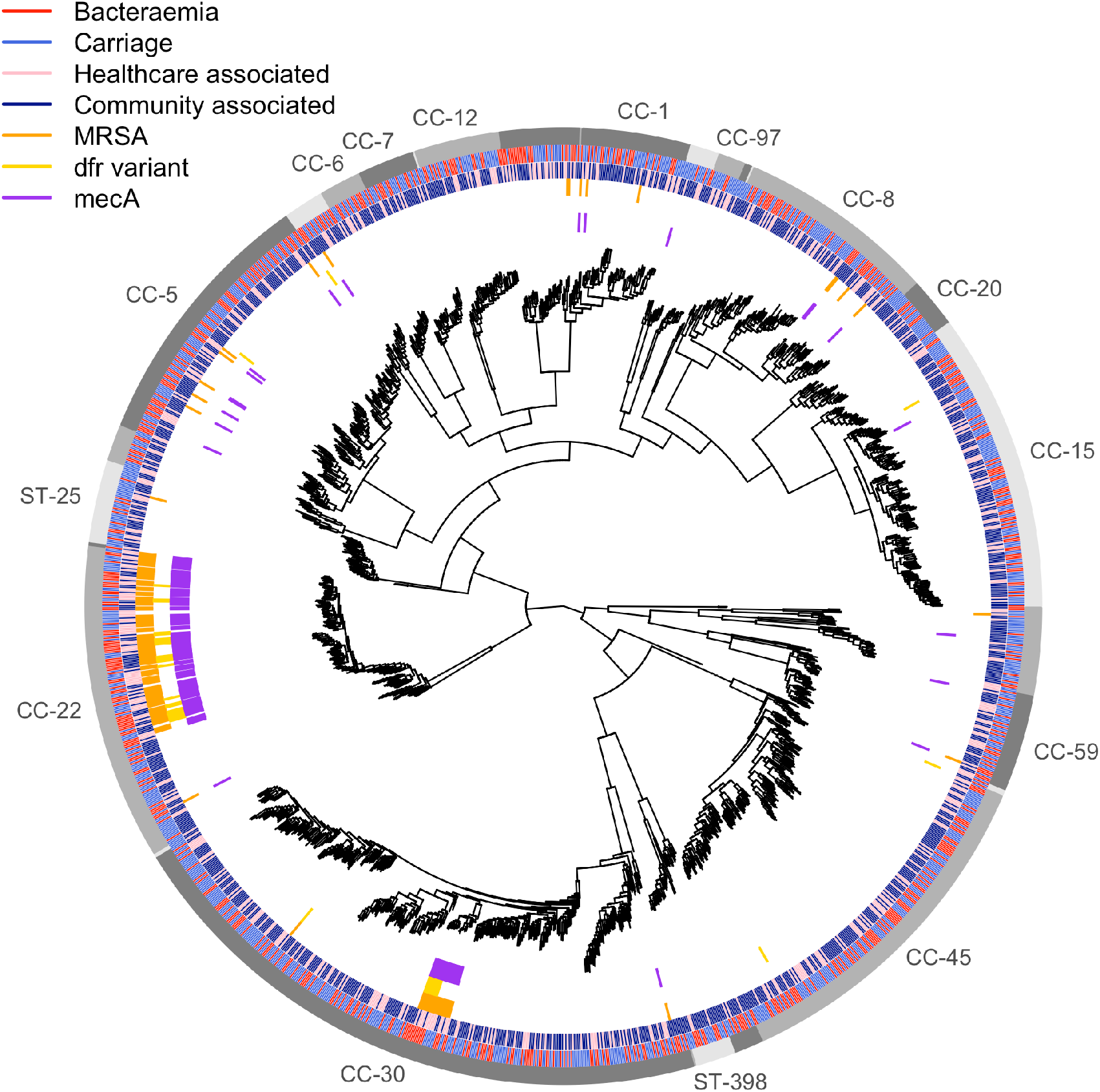
Presence of kmers that are significantly associated with carriage-vs-bacteraemia among 1610 isolates. Branch lengths have been square-root transformed to better discriminate closely related lineages. The outer ring indicates clusters with a shared lineage named by the clonal complex (or ST if only a single ST was in the cluster). The second outermost ring indicates isolate source (blue carriage, red bacteraemia). The third outermost ring indicates whether each isolate was community or hospital associated. The fourth ring indicates whether an isolate was phenotypically methicillin resistant (orange). The fifth ring marks the presence of kmers that encode a trimethoprim conferring variant in *dfrB* (yellow). The innermost ring indicates the presence of kmers mapping to the sequence of *mecA* (SAR0039).

**Figure S7:**
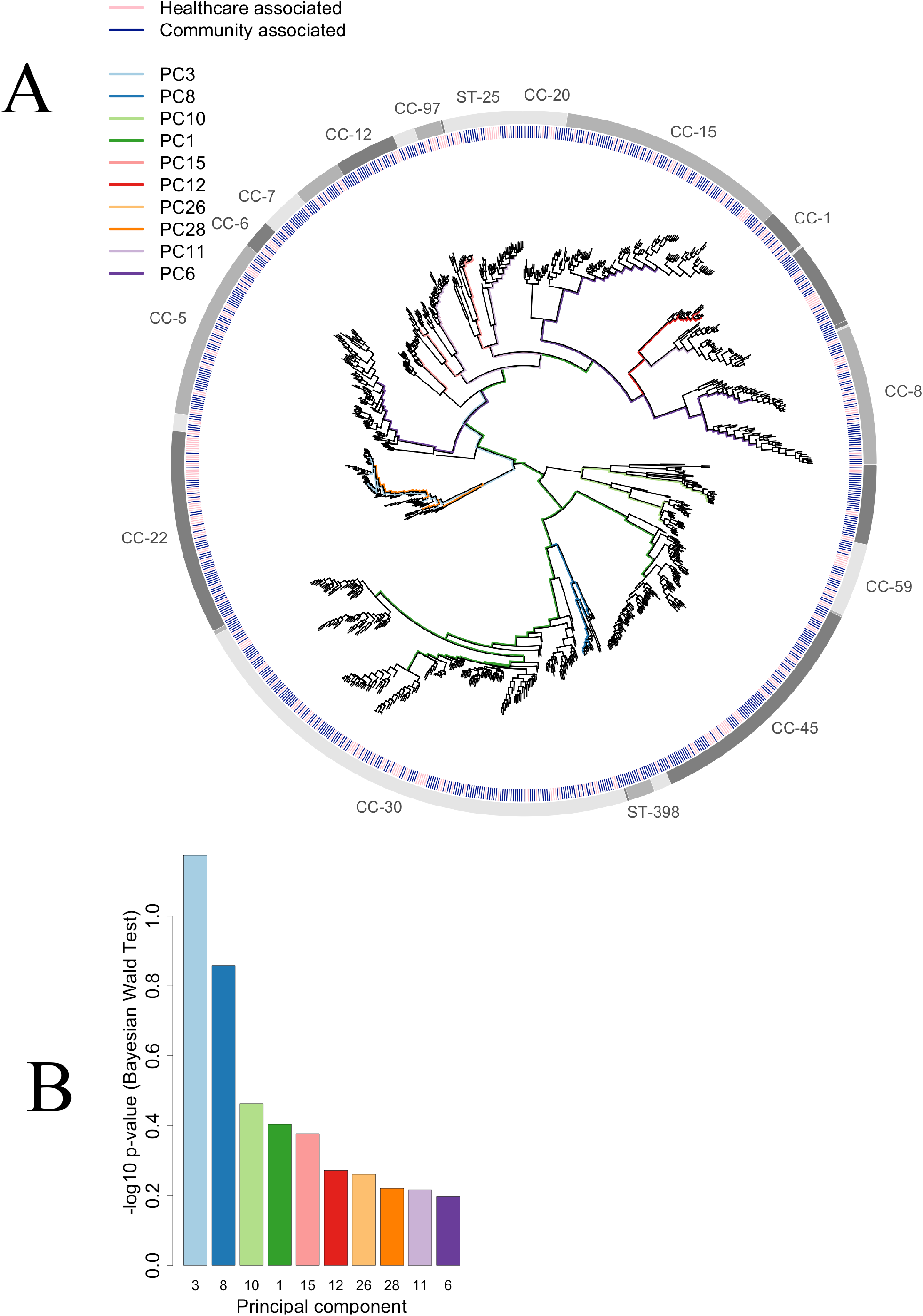
Association of Principal Components with CA-vs-HA carriage. **(A)** The branches corresponding to the 10 lineages most significantly associated with CA-vs-HA carriage are coloured on a maximum likelihood phylogeny of the study isolates. Branch lengths have been square-root transformed to discriminate closely related lineages. Clonal complexes are denoted in the outermost ring. The second outermost ring indicates isolate source (blue community, pink hospital) **(B)** Significance (negative log_10_ *p*-values) of the 10 most significantly associated PCs. A Bonferroni corrected threshold for significance is 10^−4.3^.

**Figure S8:**
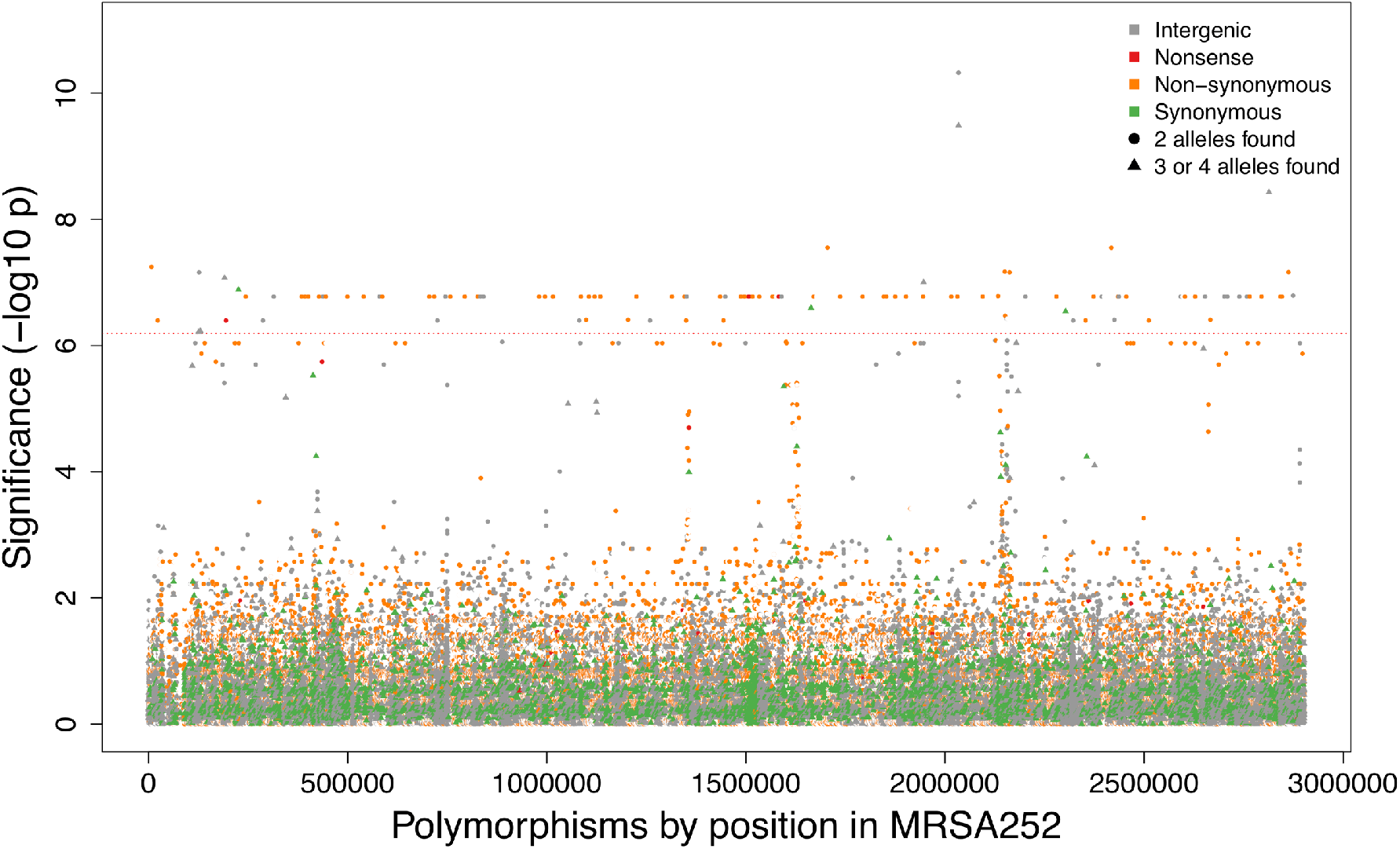
Manhattan plot of SNP associations with CA-vs-HA *S. aureus* carriage. The significance of each SNP (–log_10_ *P* value) is plotted according to location on the 2.9MB MRSA252 reference genome, with control for population structure. SNPs are coloured according to their predicted effect on protein, and shaped according to the number of alleles found at that site in the study set. We applied Bonferroni correction to the intended 5% false positive rate given the number of SNP phylopatterns to obtain a genome-wide significance threshold of 10^−6.2^ (red dotted line).

**Figure S9:**
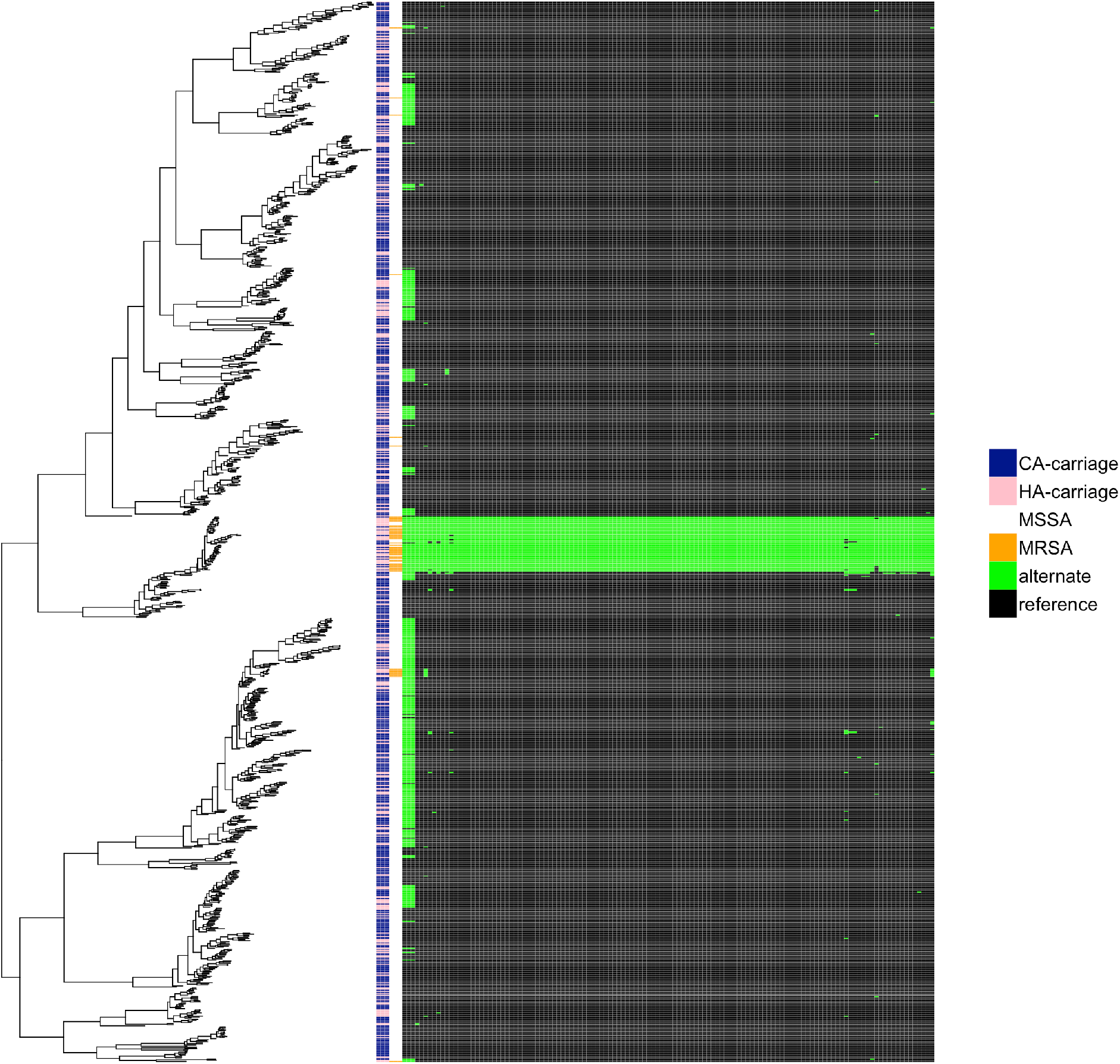
Predicted presence or absence of SNPs significantly associated with HA-vs-CA carriage among the carriage population. A maximum likelihood phylogeny of 984 carriage isolates is plotted with a matrix representing phenotypes concatenated across the most significant SNP genotypes, in order of the significance of association. A maximum likelihood tree of carriage isolates is annotated with the predicted origin of carriage (pink HA, blue CA), the presence of MRSA (orange). The remaining columns represent the presence of the reference (black) or alternate (green) allele for all significant SNPS (after imputation) which were biallelic in the study population.

**Figure S10:**
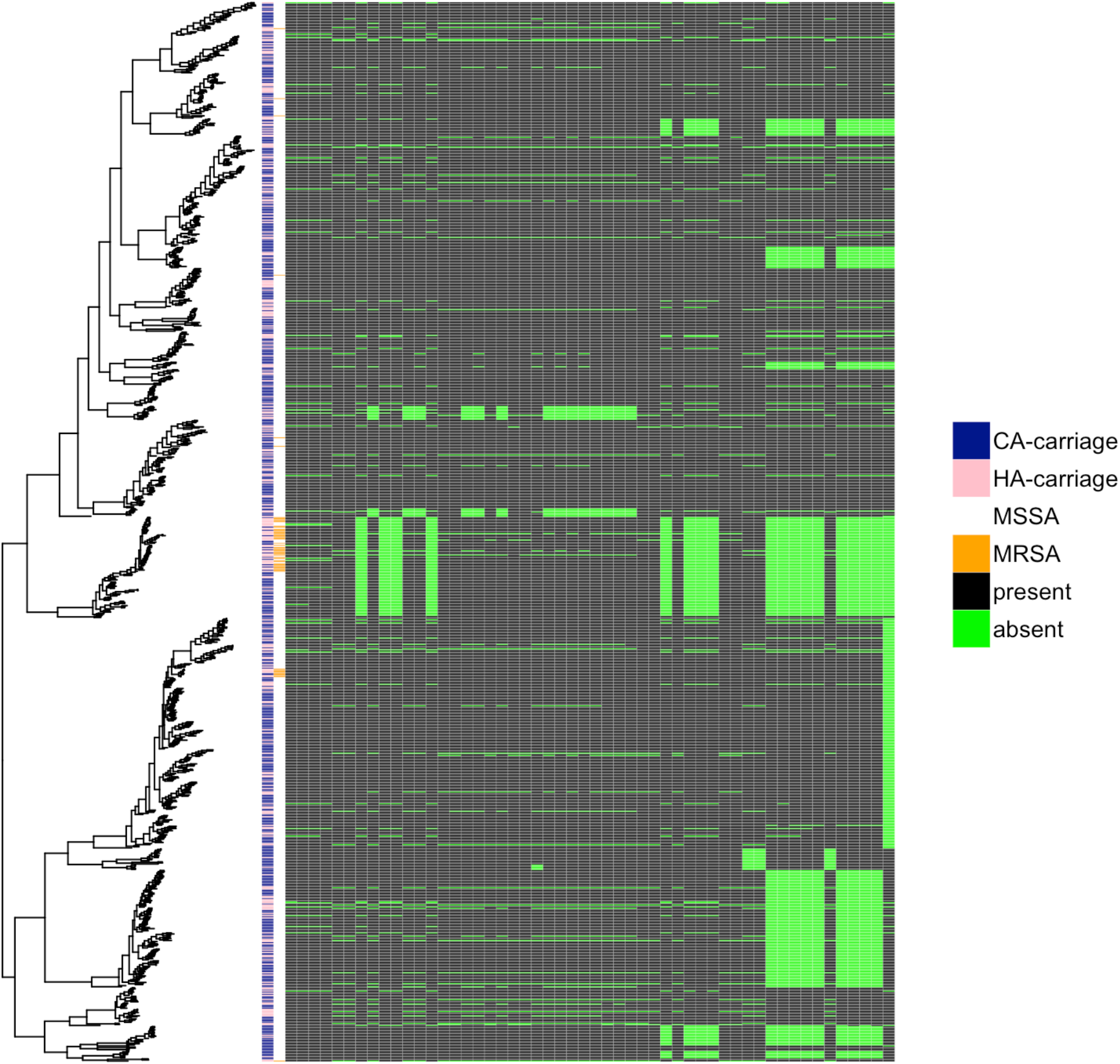
Presence of significant kmers in the population of CA and HA carriage. A maximum likelihood tree of carriage isolates is annotated with the predicted origin of carriage (pink HA, blue CA), the presence of MRSA (orange) and the presence (black) or absence (green) of each pattern of kmers mapping to *prsA*, in decreasing order of statistical significance left to right.

**Figure S11:**
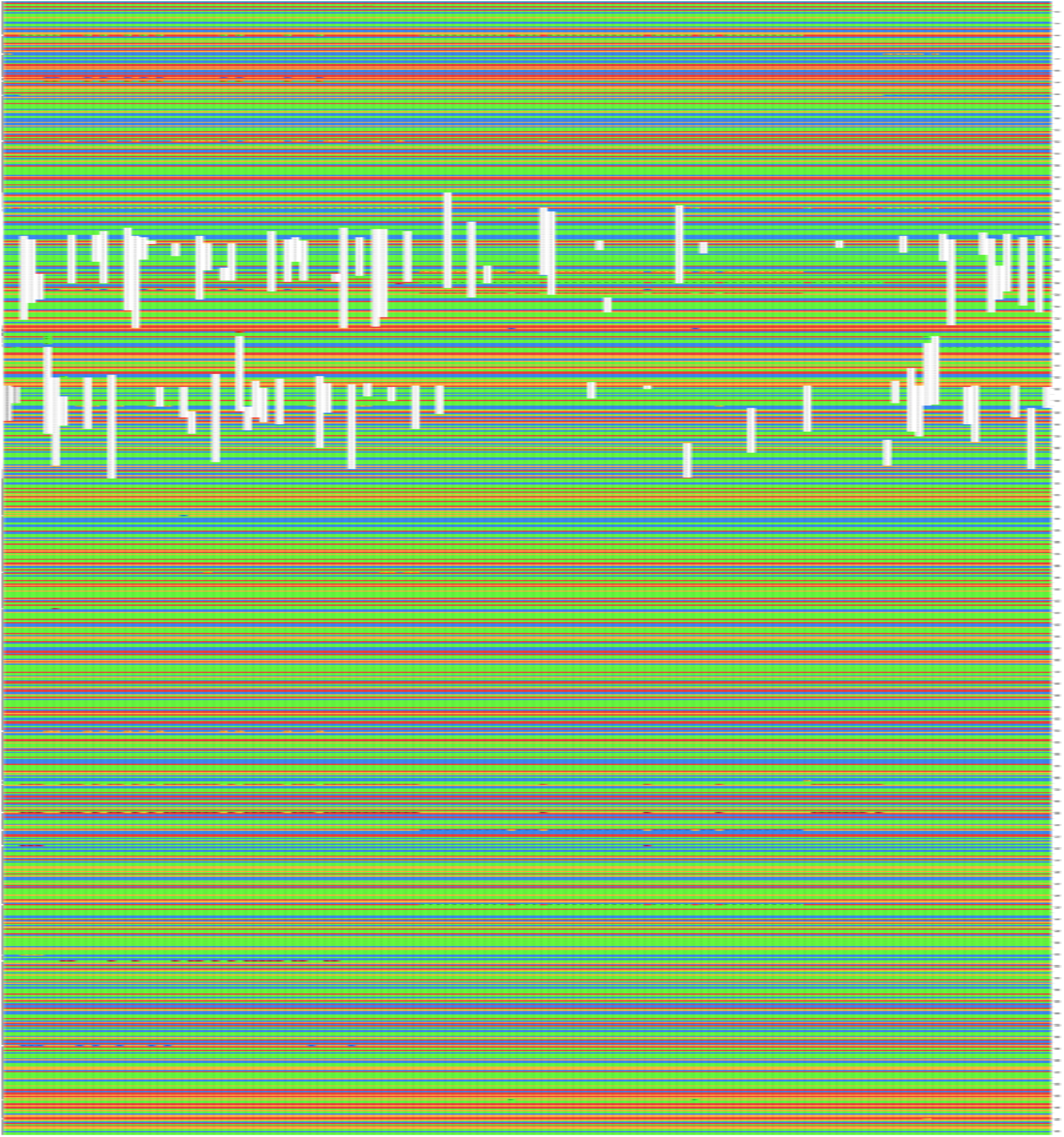
Predicted sequence of *prsA* in 131 HA-carriage genomes. The predicted sequences from 131 HA carriage genomes with the lowest number of significant *prsA* kmers found were aligned against the reference (NC_002952.2:2011797-2012759), and are displayed here aligned vertically and coloured by nucleotide (A green, C orange, G red, T blue, null – white) A plurality of variation is found, including SNPs and short deletions up to 87bp. Deletions are found across bases 164-406; the absence of kmers mapping to bases 177-394 was associated with HA carriage (Figure 3B).

**Table S1:** Focus of infection for cases (n=1017) of *Staphylococcus aureus* bacteraemia included, and compared with focus of infection reported in a large cohort of SAB in the UK.^6^

| Focus | n | % | UKCIRG (%) |
| --- | --- | --- | --- |
| Central venous catheter | 101 | 10.9 | 18.2 |
| Peripheral venous catheter | 47 | 5.1 | 5.8 |
| Endocarditis | 42 | 4.7 | 5.8 |
| Soft tissue | 244 | 26.4 | 19.5 |
| Respiratory | 52 | 5.6 | 3.7 |
| Urinary | 35 | 3.8 | (NA) |
| Thrombus | 11 | 1.2 | (NA) |
| Vascular implant | 17 | 1.8 | (NA) |
| Other | 134 | 14.5 | 13.6 |
| Not established | 235 | 25.5 | 18.8 |
| (Missing data) | 94 | 9.2% |  |

**Table S2:** Mortality at 7, 30 and 90 days, total (percentage). Differences between centres and 7, 30 and 90 days were not statistically significant (*p*=0.25, p=0.6 andp=0.9, χ^2^ test with 2 degrees of fredom)

| Centre | All cause mortality<br>by 7 days | All cause mortality<br>by 30 days | All cause mortality<br>by 90 days | Mortality data<br>missing |
| --- | --- | --- | --- | --- |
| Oxford | 94 | 180 | 228 | 6/670 (0.9) |
| N=664 | (14.1) | (27.1) | (34.3) |  |
| Brighton | 15 | 27 | 38 | 70/187 (37.4) |
| N=117 | (12.8) | (23.1) | (32.5) |  |
| Plymouth | 14 | 38 | 54 | 6/160 (3.7) |
| N=154 | (9.1) | (24.7) | (35.1) |  |
| **All** | 123 | 245 | 320 | 92/1017 (9.0) |
| **N = 925** | (13.3) | (26.5) | (34.6) |  |

Table S3: SNPs showing significant association with HA-vs-CA carriage

Table S4: Kmers showing significant association with HA-vs-CA carriage

